# Patterns of SARS-CoV-2 exposure and mortality suggest endemic infections, in addition to space and population factors, shape dynamics across countries

**DOI:** 10.1101/2021.07.12.21260394

**Authors:** Nicholas M. Fountain-Jones, Luke Yates, Emily Flies, Andrew Flies, Scott Carver, Michael Charleston

## Abstract

Some countries have been crippled by the severe acute respiratory syndrome coronavirus 2 (SARS-CoV-2) pandemic while others have emerged with few infections and fatalities; the factors underscoring this macro-epidemiological variation is one of the mysteries of this global catastrophe. Variation in immune responses influence SARS-CoV-2 transmission and mortality, and factors shaping this variation at the country level, in addition to other socio-ecological drivers, may be important. Here, we construct spatially explicit Bayesian models that combine data on prevalence of endemic diseases and other socio-ecological characteristics to quantify patterns of confirmed deaths and cases across the globe before mass vaccination. We find that the prevalence of parasitic worms, human immunodeficiency virus and malaria play a surprisingly important role in predicting country-level SARS-CoV-2 patterns. When combined with factors such as population density, our models predict 63% (56-67) and 76% (69-81) of confirmed cases and deaths among countries, respectively. While our findings at this macro-scale are necessarily associative, they highlight a need for studies to consider factors, such as infection by other pathogens, on global SARS-CoV-2 dynamics. These relationships are vital for developing countries that already have the highest burden of endemic disease and are becoming the most affected by the SARS-CoV-2 pandemic.

## Introduction

Since the severe acute respiratory syndrome coronavirus 2 (SARS-CoV-2) pandemic began in December 2019, there has been intense interest in the factors shaping disease patterns. The global patterns of cases and deaths have been relatively stable throughout the pandemic (excluding the recent dramatic rise in cases and deaths in India), with significant differences between countries that are not likely just a product of surveillance [1] or government response strategies. For example, countries in Africa with high population density and limited hospital capacity have had relatively few deaths and cases that are not simply a product of less surveillance [2,3] or a strong government response. Model estimates of 600,000 deaths [4] across the continent are not close to being realised (65 602 deaths, December 2020, [5]), and hospitals are reporting relatively low numbers of people with coronavirus disease 2019 (COVID-19) symptoms. There is substantial variation within Africa, with countries such as South Africa carrying a much higher disease burden than many of its northern neighbours [5]. Macroecological models linking environmental and climate data to outbreak data have found limited support for environmental factors shaping epidemiological dynamics [6,7]. For example, contrary to model predictions, countries in South America, such as Brazil, with similar equatorial climates, have suffered large numbers of fatalities and remain hotspots of transmission in 2020/21. Here we test a hypothesis that pathogen exposure history and population structure are more important factors driving these disparate global disease dynamics at the national scale. We predicted that countries with a high prevalence and diversity of endemic human pathogens [8], may be partially protected from the SARS-CoV-2 pandemic.

Differences in immune responses to SARS-CoV-2 infection are important for individual disease and mortality. Severe cases of COVID-19 are generally the result of an overactive immune response that results in patient mortality via acute respiratory distress syndrome [9]. The Hygiene/Old Friends Hypothesis (hereafter ‘old friends hypothesis’) is one mechanism that may underlie this overactive immune response [10]. The old friends hypothesis posits that previous exposure or coinfection by microbes with a long evolutionary relationship with humans can dampen the immune response to more novel pathogens. Prior exposure to intestinal parasitic worms such as *Ascaris lumbricoides* (ascariasis), hookworm (*Ancylostoma duodenale* or *Necator americanus*), and whipworm (*Trichuris trichiura*) are potential candidate ‘old friends’ that may mediate inflammation and the immune responses to SARS-CoV-2 [10,11]. Infection by parasitic worms is often asymptomatic but they are associated with immunosuppression and may skew cytokine response profiles that can reduce inflammation in patients exposed to SARS-CoV-2 [10–12]. However, it may not be just ‘old friends’ that may offer some protection from SARS-CoV-2. Researchers noted early in the pandemic that countries with endemic malaria (*Plasmodium* sp.) had lower than anticipated numbers of SARS-CoV-2 cases and deaths [13]. Subsequently, research found that healthcare workers coinfected with malaria recovered from SARS-CoV-2 faster than those not infected with the parasite [13].

Conversely, pathogens such as human immunodeficiency virus (HIV) [14], acute hepatitis [15] and schistosomiasis [16] may lead to increased susceptibility and the likelihood of mortality from SARS-CoV-2. Initially, HIV was not considered a risk factor for SARS-CoV-2 complications. In Europe and the United States, SARS-CoV-2 incidence was lower in HIV patients than the rest of the population as they were more likely to take precautionary measures [17]. However, there is increasing evidence that individuals infected with HIV who are untreated or have low CD4 cell counts are more at risk of mortality [14,18]. Schistosomiasis endemicity may also be associated with negative COVID-19 outcomes. For example, case numbers are higher in African regions where schistosomiasis is endemic than non-endemic areas [16]. Moreover, it may not be exposure to one particular pathogen that increases or decreases the risk of SARS-CoV-2 infection or mortality, but the diversity of pathogens endemic in a country that is important in shaping immune responses and the overall SARS CoV-2 pattern (i.e., exposure to a diversity of pathogens may damp down immune responses [19]) or prevent disease through immune memory. Given the potential importance of exposure history and coinfection, the exclusion of these variables from most macroecological models of SARS-CoV-2 is an important oversight.

The ability to distinguish meaningful relationships from spatial artefacts among macroecological predictors of SARS-CoV-2 dynamics is important. For example, national pathogen prevalence and diversity is tied to climate and economic variables. Countries closer in space may have similar numbers of cases due to the increased chances of exporting the virus across borders [20]. International air travel is also known to facilitate SARS-CoV-2 spread between countries [21], and countries with high connectivity with other countries may have increased numbers of introduction events. Further, while sharing health resources among neighbouring communities could reduce mortality rates, countries with health systems overwhelmed with COVID cases have few resources to share. High connectivity between neighbouring countries could also facilitate the spread of the virus and increase mortality. Age is also a well-known risk factor, so countries with younger populations are likely to have experienced reduced mortality than countries with higher mean ages [e.g., 1]. Country average age may also impact cases as there is a well-known bias in testing older demographics [22,23]. Economic variables such as gross domestic product (GDP) and health spending can influence the number of detected cases and deaths [24] as well as many possible predicting factors like exposure history and population demographics.

Here we use global data to construct Bayesian generalised additive models (GAMs) to untangle the connections between factors shaping the number of SARS-CoV-2 cases and deaths across countries. Results from our macroecological models can inform mechanistic research (i.e., hypothesis *generating*) to help understand varying vulnerabilities to this pressing global disaster.

## Methods

### Data retrieval

We extracted confirmed cases and confirmed deaths (per million people, hereafter ‘cases’ and ‘deaths’) for each country from the World Health Organization (WHO) on the 26th of February 2021 before widespread vaccination (https://covid19.who.int/). In addition, we accessed the number of tests per country (per thousand people) for the same period [25]. We also used the WHO transmission classification scheme (i.e., countries with community transmission, clusters of cases only, sporadic cases and no cases) to account for differences in control success.

We downloaded the mean estimate of prevalence of 14 endemic diseases in each country per country from the Institute for Health Metrics and Evaluation (IHME) GBD (Global Burden of Disease) database (see Table S1). Using this prevalence data, we calculated endemic pathogen diversity using the inverse Simpson’s metric [26]. To measure overall infectious disease burden, we also extracted estimates of years of life lost (YLLs) for infectious diseases combined in each country in 2019. Infectious diseases with an overall maximum prevalence of <10% (in 2019) were excluded from the models as they are not likely to have a large effect on SARS-CoV-2 dynamics (Gonococcal infection, Chlamydial infection, acute hepatitis, syphilis, Leishmaniasis, measles, leprosy & Trichomoniasis). The remaining seven infectious diseases were retained in our statistical models (HIV, malaria, genital herpes, lymphatic filariasis, Ascariasis. Hookworm disease, Trichuriasis & Schistosomiasis).

Additional socioecological variables were extracted for each country from the World Bank dataset (https://data.worldbank.org/) including 5-year averages (2015-2019) for per capita GDP (in current USD), percent of the population living in urban areas (hereafter ‘percent urban’), and per-capita health care expenditure (hereafter ‘health spending’). Health spending estimates (USD) are calculated by the World Health Organization and include healthcare goods and services consumed each year. We used 2019 data for average temperature and precipitation (comparable relative humidity data was not available).

We used coordinate data (country centroids) to construct a spatial neighbourhood network to estimate how the number of cases in neighbouring countries shaped cases and deaths in each country. This variable also accounts for spatial artefacts in the data. We screened for spatial autocorrelation using Moran’s I (using the R package ‘ape’ [27]). We constructed a spatial neighbourhood network from country centroids using the ‘*spdep*’ R package [28]. We also included the 2007 air connectivity index [29] to quantify the role of country-level connectivity in predicting cases and deaths

We extracted diabetes prevalence and the average cardiovascular death rate for each country in 2017 from the ‘Our world in data’ website (https://ourworldindata.org/). We screened all variables for collinearity. We imputed missing data using the powerful *MissForest* routine [30]. In brief, *MissForests* uses the *Random forest* machine learning algorithm to predict missing values utilising the observed predictor values [see 30,31 for more details]

### Bayesian modelling

We specified a separate model for each country-level response variable: cases per million people and deaths per million people. We used the same initial set of variables for both models, except that ‘cases’ was added to the deaths model. We modelled the (count) data for both response variables using a (log-link) negative binomial distribution—an overdispersed and robust generalisation of the Poisson distribution [32]. All models were fit in a Bayesian framework using Hamiltonian Monte Carlo (HMC) methods [33] and ‘no U-turn sampling’ (NUTS, [34]), as implemented in the R package ‘brms’. We ran four chains of 9000 iterations, using weakly informative priors. To establish chain convergence, we used the rank-normalised 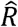 statistic 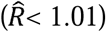 as well as visual inspection [35]. We performed model selection using approximate leave-one-out cross-validation of the posterior predictive log density (LOO, [36])—this estimates the information-theoretic, relative Kullback–Leibler discrepancy in a Bayesian setting. The selected models were validated using posterior predictive checks based on kernel density estimates and pointwise credible intervals [37].

Initially, we used LOO to compare a linear model to a smooth spline model; the latter estimates a penalised thin plate regression spline for each continuous variable, permitting non-linear dependencies (*mgcv*, [38]). Then, to select a subset of the most important variables, we applied a small penalty to the linear term(s) of the spline basis, permitting the whole term to be shrunk to zero [38]. Next, we removed all variables whose smooth functions were consistent with zero slope, and compared the performance of the reduced model to the full model using LOO. Finally, to account for heteroskedasticity, we added region-level terms to the modelled shape (variance) parameter, using LOO and posterior predictive checks to assess the merits of the added terms.

For the final selected models, we computed the conditional effects for each continuous variable. The corresponding plots show the estimated (response-scale) effect of changes in the value of a given predictor while holding all other variables at their mean. We compared the observed values to our modelled predictions, using the 95% credible interval (CI) of the posterior predictive distribution to identify countries where our model was not adequate (i.e., the observation was outside the 95% CI). Our complete workflow and data are available on github (https://github.com/nfj1380/covid19_macroecology).

## Results

Our models reveal the importance of endemic pathogen prevalence, space and demography in explaining the number of cases and deaths for countries across the globe. For each model, we used posterior predictive checks to verify that our models could adequately predict the observed data (Fig. S1), and Bayesian R^2^ estimates as a goodness-of-fit measure (case model: 0.63 (credible interval (CI): 0.56 -0.67, death model: 0.76 (CI: 0.69-0.81)). However, the pathogens and population characteristics that had the most predictive power, and were thus retained in the final model, differed between the cases and deaths models. Variables retained in the case model included the country-level prevalence of HIV, Ascariasis, malaria, and percent urban (Fig. 1), whereas hookworm disease, population density and mean population age were more important in the deaths model (Fig. 2). The spatial neighbourhood network was also important in both models. Average temperature, average rainfall, GDP and per capita health care expenditure were not important in either model. Including region in the error term also improved model performance overall, indicating some impact of region in the prediction of SARS-CoV-2 dynamics. See Table S2 for LOO model comparison results.

**Fig 1:**
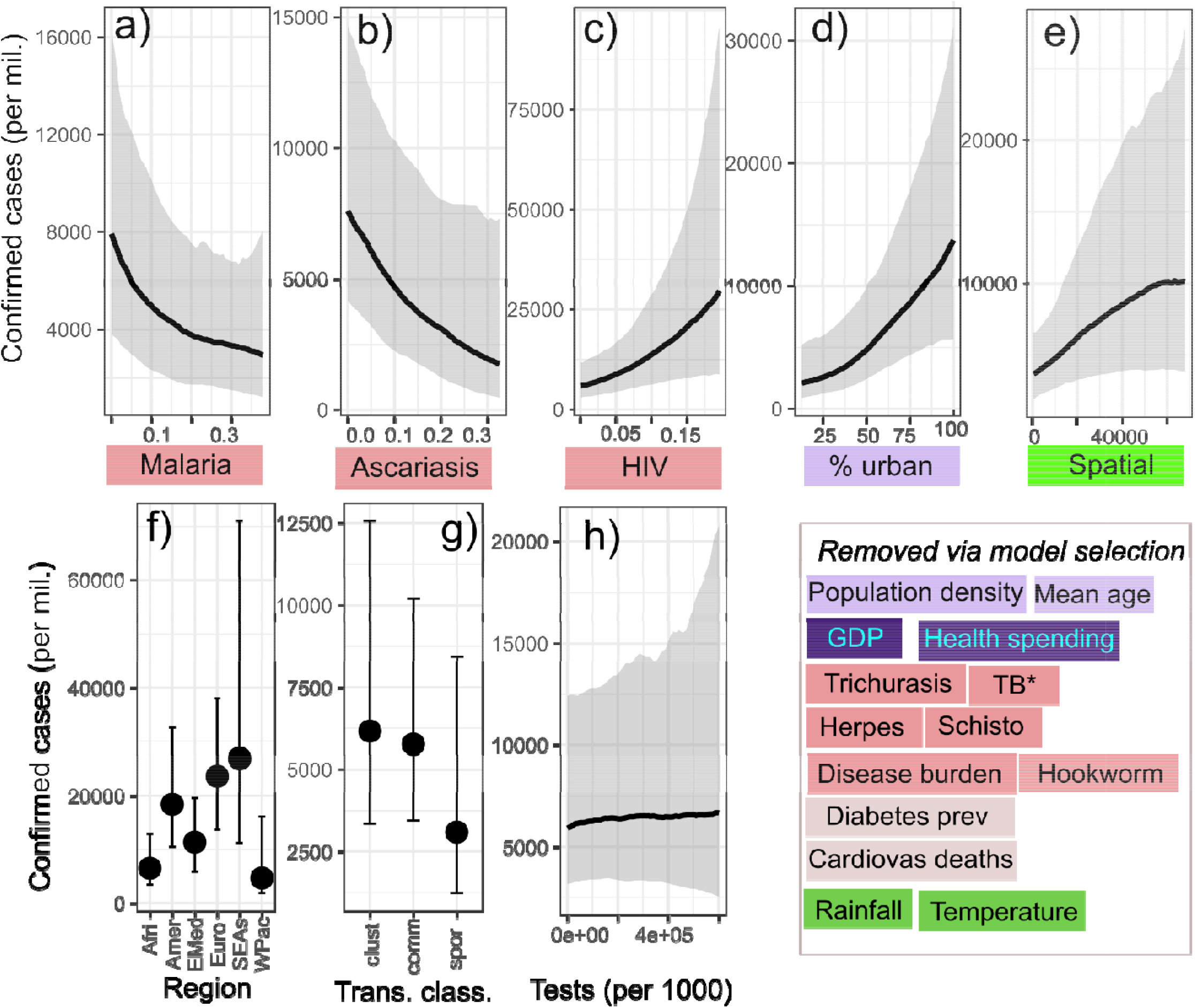
Conditional effects plots for the cases model. Black lines denote the posterior means and the associated intervals (or grey shading) denote the 95% credible intervals. a-h) plots for predictors included in the model. Predictors excluded from the model are provided in the lower right box. Predictors are colour-coded based on variable type (dark blue = associated with a country’s economic capacity, light red = country level pathogen prevalence estimate, light grey = estimated prevalence of non-infectious disease, green = climate variable, yellow = spatial variable, light blue = population characteristic. Predictors without coloured boxes are variables included in the model to account for biases in the data. *: estimates of cases in neighbouring countries. TB: Tuberculosis prevalence, HIV: Human immunodeficiency virus. L.filariasis = lymphatic filariasis. Regions (per WHO): Afri: Africa, Amer = Americas, EMed = Eastern Mediterranean, SEAs = Southeast Asia, WPac = Western Pacific. Clust = Clusters of cases identified in a country, Comm = community transmission, spor = sporadic cases detected. See Table S1 for variable details, Fig. 3 for the spatial distribution of model variables and Fig. S2 for correlations between predictors.

**Fig 2:**
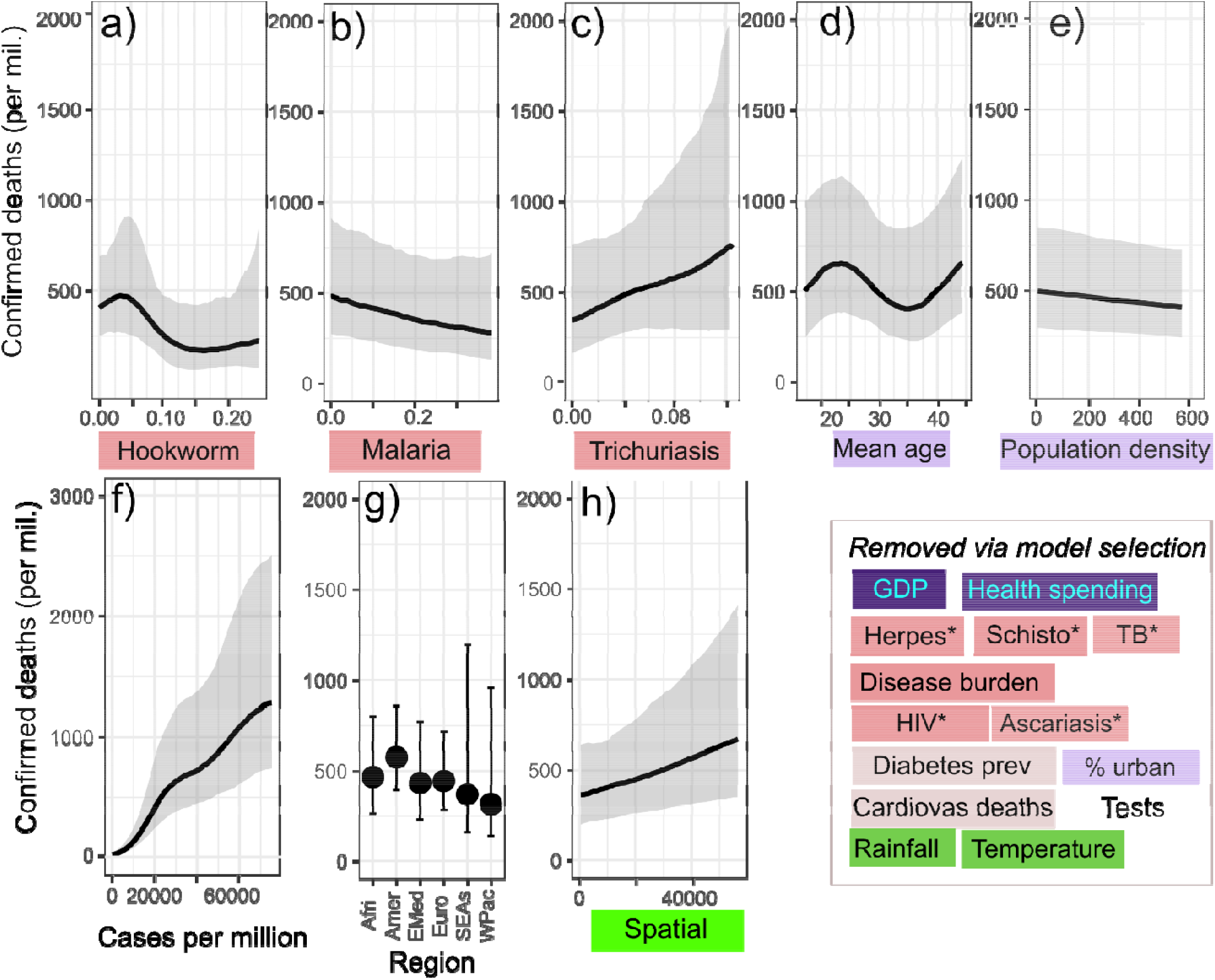
Conditional effects plots for the deaths model. Confidence intervals represent 95% credible intervals based on the posterior distribution. Predictors excluded from the model are provided in the lower right box. a-h) plots for predictors included in the model. Predictors are colour coded based on variable type (dark blue = associated with a countries economic capacity, light red = country level pathogen prevalence estimate, light grey= estimated prevalence of non-infectious disease, green = climate variable, yellow = spatial variable, light blue = population characteristic. g) shows the region-level intercepts which can be viewed as a base line value before the modelled effects of covariates are included. The actual number of (modelled) deaths will depend on the covariate values in each country, so higher intercept values will not necessarily translate to higher deaths. Predictors without coloured boxes are variables included in the model to account for biases in the data. *: estimates of cases in neighbouring countries. TB: Tuberculosis prevalence, HIV: Human immunodeficiency virus. L.filiariasis = lymphatic filariasis. Regions (per WHO): Afri: Africa, Amer = Americas, EMed = Eastern Mediterranean, SEAs = South-East Asia, WPac = Western Pacific. See Table S1 for variable details, Fig. 3 for the spatial distribution of model variables and Fig. S2 for correlations between predictors.

We excluded GDP as it was highly collinear (ρ >0.7, see Fig. S2) with health spending. We excluded air connectivity as it was highly positively correlated with mean age and the spatial network (i.e., countries with high air connectivity were close to each other and had a similar mean age). We also excluded pathogen diversity as it was strongly negatively correlated with mean age (i.e., countries with high pathogen diversity had lower mean ages), and the proportion older than 65 years in the population as it was positively associated with mean age (Fig, S2).

Plotting the conditional effects of each predictor (while holding all other predictors at their mean value) revealed that prevalence estimates of other pathogens and population characteristics had divergent modelled effects on cases and deaths in each country (Figs. 1 & 2). For example, there were negative, approximately linear, relationships between SARS-CoV-2 cases and both malaria and Ascariasis (Figs. 1a/1b). Countries with an estimated prevalence > 0.2 (20%) of both endemic pathogens have approximately half the mean number of SARS-CoV-2 cases of countries with low prevalence (Figs. 1a/1b). Central Africa was the only region where estimates of malaria prevalence exceeded 0.2 (20%). In contrast, ascariasis prevalence was relatively high on the sub-continent, South-East Asia and central and western Africa (Fig. 3d). We also found a non-linear relationship between hookworm infection and SARS-CoV-2 deaths. For example, countries with > 0.1 prevalence (exclusively in Central Africa, Fig. 3e) have approximately half the SARS-CoV-2 deaths (Fig. 2a). Countries with high malaria prevalence also had fewer SARS-CoV-2 deaths but this effect was minor (Fig. 2b). In contrast, we detected positive, approximately linear, relationships between SARS-CoV-2 cases and HIV prevalence (Fig. 1c) and between SARS-CoV-2 deaths and Trichuriasis prevalence (Fig. 2c).

**Fig. 3:**
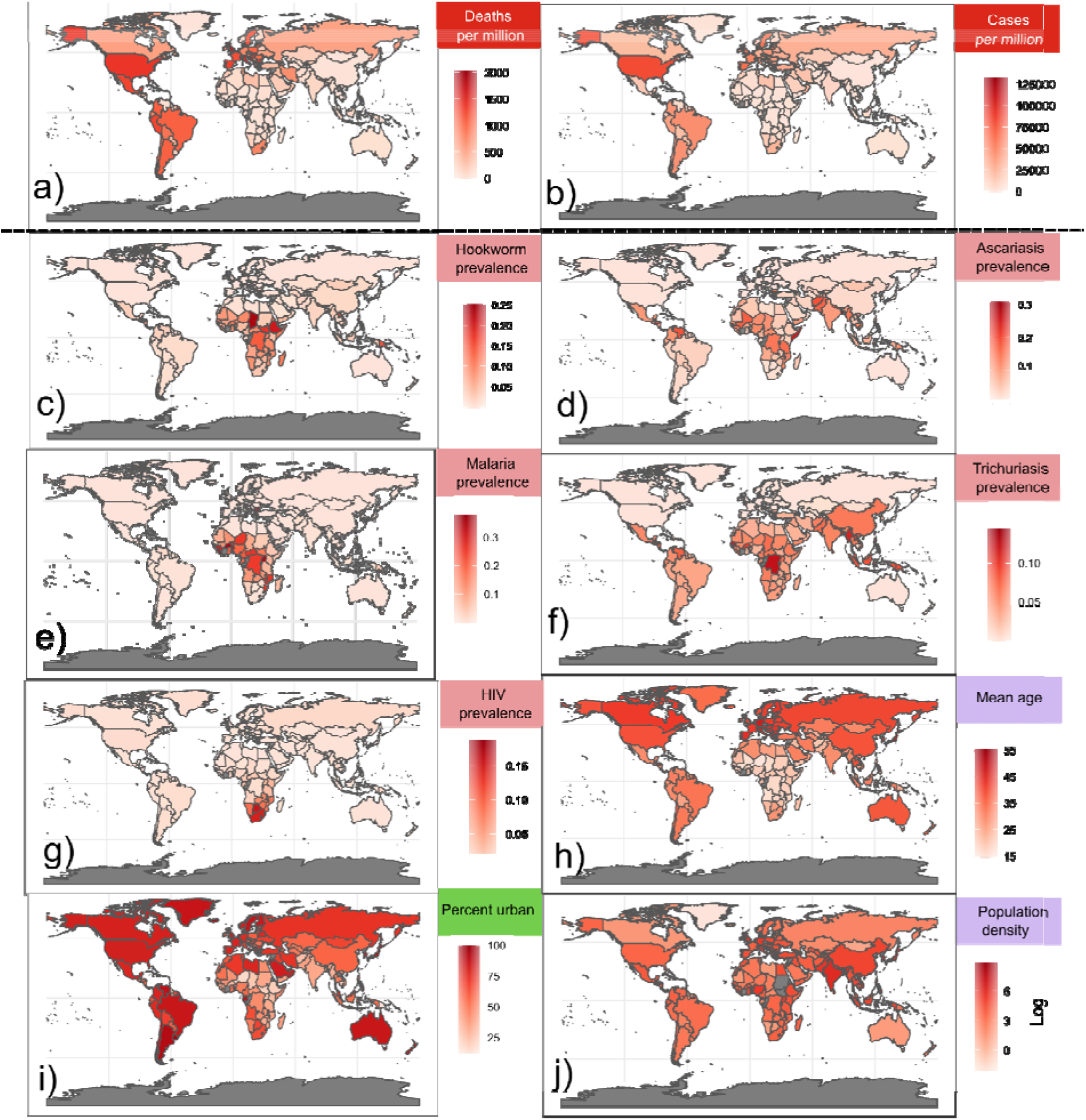
Maps showing the global patterns of each set of predictors used in our model. See Table S1 for details of each predictor. The first two maps separated by the dashed lines are the response variables (a) cases and (b) deaths, (c-j) are predictors in each respective model.

Population characteristics also had variable effects on the number of SARS-CoV-2 deaths and cases in each country. There was a positive, approximately linear, relationship between the proportion of a country’s population living in urban areas and SARS-CoV-2 cases (Fig. 1d) with this proportion lowest in central Africa (Fig. 3). In contrast, mean age had a strongly fluctuating non-linear effect on SARS-CoV-2 deaths, with deaths peaking at a mean age of 22 and 50 yrs, and minimal when the mean age was around 35 (Fig. 2d). Population density had a minor negative relationship with deaths for each country (Fig. 2e). Central Africa had the lowest mean age and the northern latitudes of the continent had some of the lowest human population density values (Fig. 3j).

There was a strong positive, approximately linear, relationship between SARS-CoV-2 cases and deaths in each country (Fig. 2f). The cases in the spatial neighbourhood also showed a positive, mostly linear fit in both models (Figs. 1e/2h). We did not find a conditional effect of estimated tests on SARS-CoV-2 cases. However, there was some variation in cases explained by region, with the Western Pacific and Africa having a lower baseline (region-level intercept) than the other regions after controlling for all other variables in the model (Fig. 1f). For the SARS-CoV-2 deaths models, regional variation of the baseline was negligible (Fig.

2g). For both models, the inclusion of region-level terms to the modelled variance (Poisson overdispersion) significantly improved predictive performance (Table S2); these terms quantify the differing variance of the (modelled) distribution of the data between regions (Fig S3).

Overall, our models predicted the number of SARS-CoV-2 cases and deaths well, with model estimates from only eight countries out of the 181 included being outside our 95% credible interval (Fig. 4). The uncertainty of the modelled predictions varied by region and response (Fig. 4). European estimates of SARS-CoV-2 cases and deaths had the highest precision (i.e., smallest credible intervals) in our models and estimates with the lowest precision included the Western Pacific (e.g., China, Australia, Cambodia) and South-East Asian WHO regions (Fig. 4, Fig. S4). The precision of our model estimates of cases for the African region were comparable to Europe, the Americas and Eastern Mediterranean (Fig. 4, Fig. S4). Tanzania was the only country in which the models overestimated cases and deaths. Our models also overestimated the number of SARS-CoV-2 cases in Panama, Yemen, Guam and Cabo Verde and deaths in Benin and Mexico. The only countries our models underestimate were cases in Nicaragua (see Fig. S3).

**Fig. 4:**
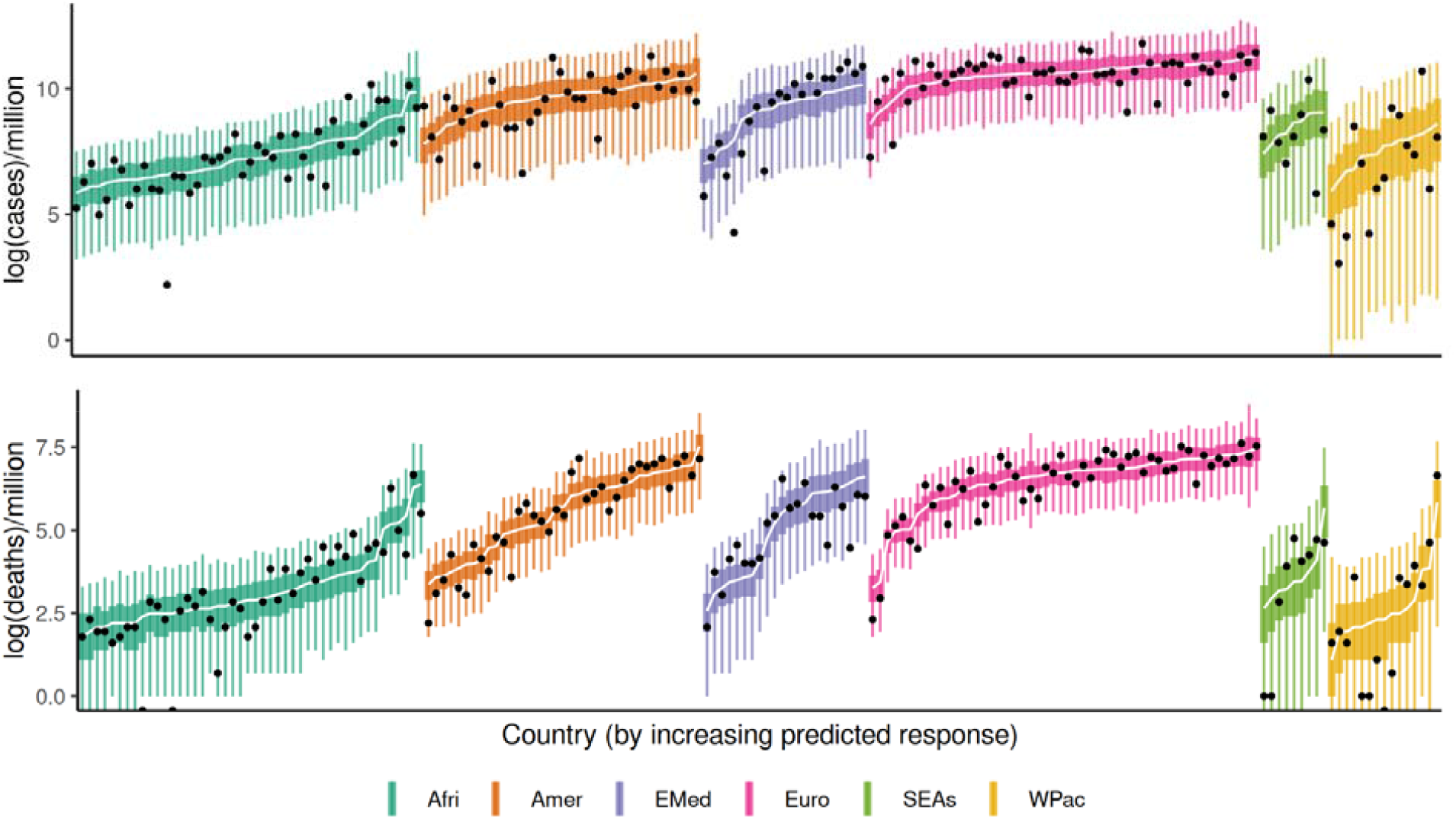
Predictive performance of our SARS-CoV-2 cases model (top panel) and SARS-CoV-2 deaths model (bottom panel) for each country. Countries are grouped by region and ordered by predicted values (lowest to highest). Black dots are the observed values for each country and the trend line is the median posterior estimate of the models. Credible intervals (CIs) are coloured by region, with the 50% and 95% CIs depicted by thick and thin lines, respectively. Afri: Africa, Amer = Americas, EMed = Eastern Mediterranean, SEAs = South-East Asia, WPac = Western Pacific. See Fig. S3 for predictions with each country labelled.

## Discussion

A mix of endemic pathogen prevalence estimates, spatial data and population characteristics predicted SARS-CoV-2 dynamics across the globe remarkably well. Our Bayesian analytical approach revealed that the percentage of people living in urban centres, the number of cases in neighbouring countries, and the prevalence of endemic pathogens all shaped the observed patterns of cases prior to widespread vaccination. For example, higher HIV prevalence was associated with higher SARS-CoV-2 case numbers, whereas increased ascariasis prevalence had the opposite effect at the country level. The number of SARS-CoV-2 cases was not the only predictor of deaths, with countries with a high prevalence of hookworm and malaria experiencing fewer SARS-CoV-2 deaths compared to those with lower hookworm and malaria prevalence. Strikingly, mean temperature, rainfall, healthcare spending and noncommunicable disease, often central to many SARS-CoV-2 macroecological models [e.g., 1,39,40], were less important in explaining the patterns of cases and deaths across countries. Over a year into the pandemic, African countries that suffer the most from the burden of endemic infectious disease have been less impacted by SARS-CoV 2, and our models point to some potential explanations. We stress that at this global scale, there is uncertainty and the relationships we observed are not necessarily mechanistic; also, the data we used are likely substantial underestimates of cases and deaths. Rather, our study highlights the need for more focussed experimental work or case-control studies to understand better the complex interplay between endemic pathogens and population characteristics in shaping the health outcomes in this global emergency.

We demonstrate that estimates of endemic pathogen prevalences are valuable predictors of both SARS-CoV-2 cases and deaths and have utility in predicting SARS-CoV-2 dynamics at the global scale. Given the likely ubiquity of coinfections of these common pathogens in large parts of the world and increasing evidence that coinfection can impact the severity of SARS-CoV-2 [11,13], it is plausible that our population-level results reflect individual-level coinfection patterns. For example, people infected by HIV often have low CD4 cell counts and can face a higher risk of severe SARS-CoV-2 infection [41], and in our models HIV was associated with higher case numbers, which in turn increases deaths. Countries in Southern Africa have the highest number of cases and deaths per million people on the continent and have the highest estimated prevalence of HIV in the world (Fig. 3). Further, there is evidence that malaria can also shape SARS-CoV-2 dynamics by reducing the severity of the disease. For example, a study of healthcare workers in India found that patient recovery from SARS-CoV-2 infection was on average eight days faster with coinfection of malaria than without malaria [13]. Lab trials have shown that there may be some cross-reactivity between *Plasmodium* specific antigens with SARS-CoV-2 antibodies that may explain reduced severity of COVID-19 and may help prevent infection [42]. Malaria prevalence was important in both case and death models, which helps to explain the observation that malaria-endemic countries tended to have fewer cases and deaths [43].

Prevalence of helminth infections were also important in both SARS-CoV-2 case and death models, but the nature of the relationship varied (e.g., countries with high trichuriasis prevalence had higher numbers of deaths, whereas the opposite was true for hookworm). Whether helminth infection can suppress or increase SARS-CoV-2 severity has been debated [10,11,44]. Helminths are hypothesised to increase COVID-19 mortality as worm infections can elevate maladaptive interleukin-4 (IL-4) and IL-10 cytokines associated with severe cases of COVID-19 [11]. However, modulation is also possible as helminth infections may reduce the likelihood that the patient suffers from metabolic diseases such as diabetes which are known risk factors for COVID-19 mortality [44]. Alternatively, helminth antigens may suppress inflammation and favour the production of immunoregulatory or anti-inflammatory cytokines [10–12]. However, there is epidemiological information that does not support the modulatory effects of helminth infections. Areas of the Amazon in Brazil, for example, have much higher COVID-19 death rates compared to the rest of the country, but also have high prevalence of ascariasis (>85%) [45]. In our global models, ascariasis prevalence generally decreased cases and in doing so, potentially reduced deaths. Trichuriasis infection, estimated to be at a similar prevalence to ascariasis in the Amazon [46], in contrast, was associated with increased cases in our model. Interestingly though, hookworm is not endemic in the Americas [45] and we found a more direct effect of the prevalence of this helminth, reducing the number of deaths attributed to SARS-CoV-2. Heterogeneity in the relationships between helminth species and other pathogens such as malaria or tuberculosis in coinfections are well understood [46,47] so it is not surprising that we see this for SARS-CoV-2. Examining regional patterns in helminth coinfections as well as focussed case-control studies will likely further clarify the nature of these potentially complicated relationships.

Population characteristics and spatial patterns were also important in our models, in particular, cases were higher in more urban nations where a higher percent of the national population lives in cities. This outcome mirrors findings from country-level analyses early in the pandemic (April 2020) [24]. Cities have a higher population density and a more transient population than surrounding areas, making them potential incubators for infectious disease and contributing to urban-associated disease patterns [48]. But disease dynamics in cities are complex: better access to socioeconomic and medical resources mean urban residents often have a higher life expectancy and lower overall disease burden than rural counterparts [49]. Even amongst infectious diseases, prevalence tends to decrease as nations become more urban, likely due to simultaneous increases in sanitation, wealth and healthcare access [50]. Importantly though, cities are also travel hubs, making them a gateway for disease incursions during a pandemic [51]. SARS-CoV-2 is a directly-transmitted pathogen that can remain active on surfaces for several days [52]. Under such circumstances, the benefits of living in cities that are present for other infectious diseases (sanitation, access to economic and medical resources) appear to be outweighed by the facilitated SARS-CoV-2.

Interestingly we found a weak pattern of a mean age on SARS-CoV-2 cases and deaths, unlike what has been found in other smaller scale analyses [53]. Other country-level models have used the proportion of population >70 as a predictor in similar models, but we found that mean age was strongly correlated with proportion over 65 (ρ = 0.9). Our analysis differs from others because we have attempted to account for and model spatial autocorrelation and regional differences (i.e., cases in one country are shaped by the number of cases in neighbouring countries). The spatial neighbourhood predictor was quite strongly correlated to mean age (ρ= 0.63), and this may have accounted for mean age variation in our models (i.e., countries with similar mean ages are neighbours with similar numbers of cases). Similar correlations have been detected in previous macroecological analyses [24]. Older adults are well known to suffer the highest mortality rate following infection with SARS-CoV-2 [e.g., 54]. In no way do our models suggest that age is not a risk factor for mortality, but rather at this scale, the effect is difficult to distinguish from other spatial patterns. Even with our ability to travel large distances rapidly, patterns of SARS CoV-2 spread are constrained by geography at regional scales [55–57]. At our global scale, Moran’s I analysis indicated that spatial autocorrelation was a feature of this dataset and controlling for it was important to help interpret the other predictors in our models.

As with any model at a global scale, there are important limitations. For example, our models cannot capture important regional differences and cannot directly infer individual-level mechanisms. Moreover, our models utilise WHO confirmed case and death data that are likely considerable underestimates of the true scale of this global emergency. Nonetheless, the overall strong relationship between cases and deaths supports the idea that modelling the case data is of utility as COVID-19 deaths are generally better reported than cases [58]. The weak relationship between cases and tests is not surprising as it has been demonstrated that regional differences can vary by country [1]. Moreover, while we assembled a diverse set of predictors, this dataset is not comprehensive. We aimed to maximise the number of countries we included in the analysis without introducing large amounts of missing data. However, the predictive performance of our models was surprisingly high, and few model estimates of cases and deaths not including the observed values for each country (Fig. 4). Countries such as Tanzania, for which our model overpredicted cases and deaths, may warrant increased attention and surveillance.

Our study shows that for any global analysis of SARS-CoV-2 dynamics, the patterns of other pathogens can provide important insights. At a broad spatial scale, variations in climate are likely to be less critical than variations in contemporary and historical exposure to a diverse array of pathogens. The immune system’s role and evolved defence mechanisms of these complex associations clearly warrant more targeted investigation. As the pandemic progresses, variant dynamics are shifting and developing countries will take a greater burden of SARS-CoV-2 cases/deaths. Hypotheses generated using these types of approaches may be increasingly valuable in managing this ongoing global catastrophe and inform outbreak management in future pandemics.

## Data Availability

All data will be available on github

https://github.com/nfj1380/covid19_macroecology/tree/master/plots

## Acknowledgements

This project was supported by an Australian Research Council Discovery Project Grant (DP190102020).

## Supplementary information

**Table S1:** Summary of predictors used to construct our models.

| Predictor | Type | Data Source | Units | Notes |
| --- | --- | --- | --- | --- |
| Number of tests | S | Hassell et al (2020)<br>https://github.com/owid/covid-19-data/tree/master/public/data/testing | Tests (per thousand) |  |
| Confirmed cases^ | S | World Health Organization:<br>https://covid19.who.int/: | Cases (per million) | Used as a predictor in the death model |
| Population density | Pop | Average from 2015-19:<br>https://data.worldbank.org/ | People per km<sup>2</sup> |  |
| Mean age | Pop | Average from 2015-19:<br>https://data.worldbank.org/ | Years |  |
| Proportion<br>> 65 y.o | Pop | Average from 2015-19:<br>https://data.worldbank.org/ | % |  |
| % urban | Pop | Average from 2015-19:<br>https://data.worldbank.org/ | % |  |
| GDP | E | Average from 2015-19:<br>https://data.worldbank.org/ | Per-capita GPD |  |
| Health spending | E | Average from 2015-19:<br>https://data.worldbank.org/ | US dollars |  |
| Hospital beds | E | Average from 2015-19:<br>https://data.worldbank.org/ | Beds per-capita | Strongly correlated to health spending* |
| Rainfall | CI | World Bank (2019 average) | Millimeters (mm) |  |
| Temperature | CI | World Bank (2019 average) | Degrees centigrade (°C) |  |
| Herpes | P | Institute for Health Metrics and Evaluation (IHME) GBD database:<br>https://vizhub.healthdata.org/gbd-compare/ | Estimated prevalence |  |
| Human immunodeficiency virus (HIV) | P | Institute for Health Metrics and Evaluation (IHME) GBD database:<br>https://vizhub.healthdata.org/gbd-compare/ | Estimated prevalence |  |
| Malaria | P | Institute for Health Metrics and Evaluation (IHME) GBD database:<br>https://vizhub.healthdata.org/gbd-compare/ | Estimated prevalence |  |
| Tuberculosis (TB) | P | Institute for Health Metrics and Evaluation (IHME) GBD database:<br>https://vizhub.healthdata.org/gbd-compare/ | Estimated prevalence |  |
| Ascariasis | P | Institute for Health Metrics and Evaluation (IHME) GBD database:<br>https://vizhub.healthdata.org/gbd-compare/ | Estimated prevalence |  |
| Hookworm | P | Institute for Health Metrics and Evaluation (IHME) GBD database: https://vizhub.healthdata.org/gbd-compare/ | Estimated prevalence |  |
| Schistosomiasis | P | Institute for Health Metrics and Evaluation (IHME) GBD database: https://vizhub.healthdata.org/gbd-compare/ | Estimated prevalence |  |
| Trichuriasis | P | Institute for Health Metrics and Evaluation (IHME) GBD database: https://vizhub.healthdata.org/gbd-compare/ | Estimated prevalence |  |
| Lymphatic filariasis | P | Institute for Health Metrics and Evaluation (IHME) GBD database: https://vizhub.healthdata.org/gbd-compare/ | Estimated prevalence |  |
| Pathogen diversity | P | Data collated from IHME GBD database and then inverse Simpson's index estimated (see *Methods*) | Inverse Simpson's index | Strongly collinear with mean age |
| Infectious disease burden | P | Institute for Health Metrics and Evaluation (IHME) GBD database: https://vizhub.healthdata.org/gbd-compare/ | Years of life lost (YLLs) |  |
| Diabetes | C | Our world in data (2017 estimate): https://ourworldindata.org/ | Prevalence |  |
| Cardiovascular deaths | C | Our world in data (2017 estimate): https://ourworldindata.org/ | Prevalence |  |
| Spatial network | Sp | Network created using the centroid coordinates of each country (see *Methods*) | Cases in countries close in space |  |
| Air travel | Sp | Arvis, et al., .2011 | Air connectivity index (2011) | Strongly correlated with GDP*. |

---

**Fig. S1:**
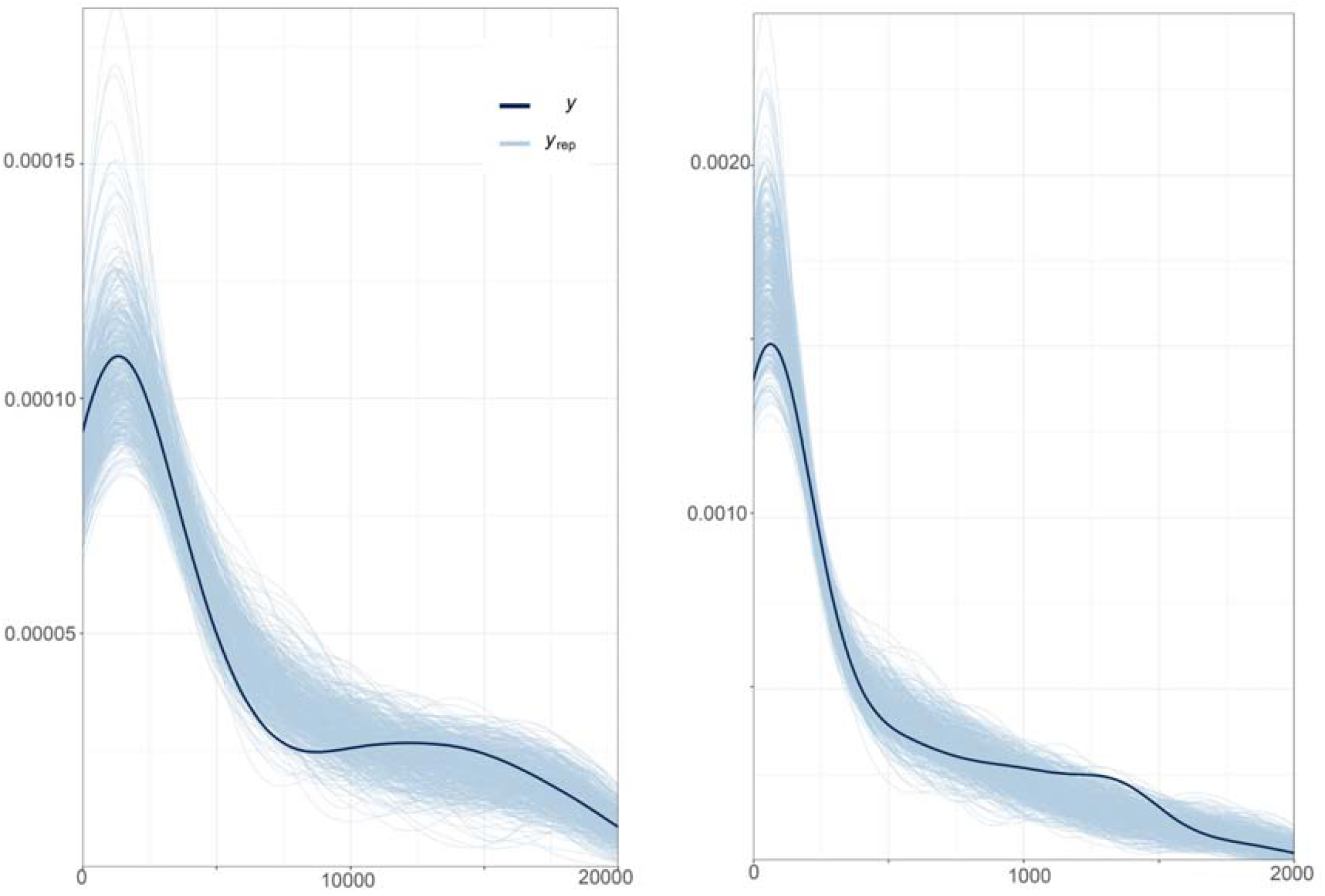
Predictive density checks showing simulated data (light blue lines) generated using parameters values drawn from the posterior distribution of the fitted models: case model (left), death model (right). The values on the x-axis are the range of response values for each model and the plotted curves are kernel density estimates of the distribution of the observed (dark blue line) and simulated data sets. These checks illustrate the ability of the models to generate plausible data.

**Fig. S2:**
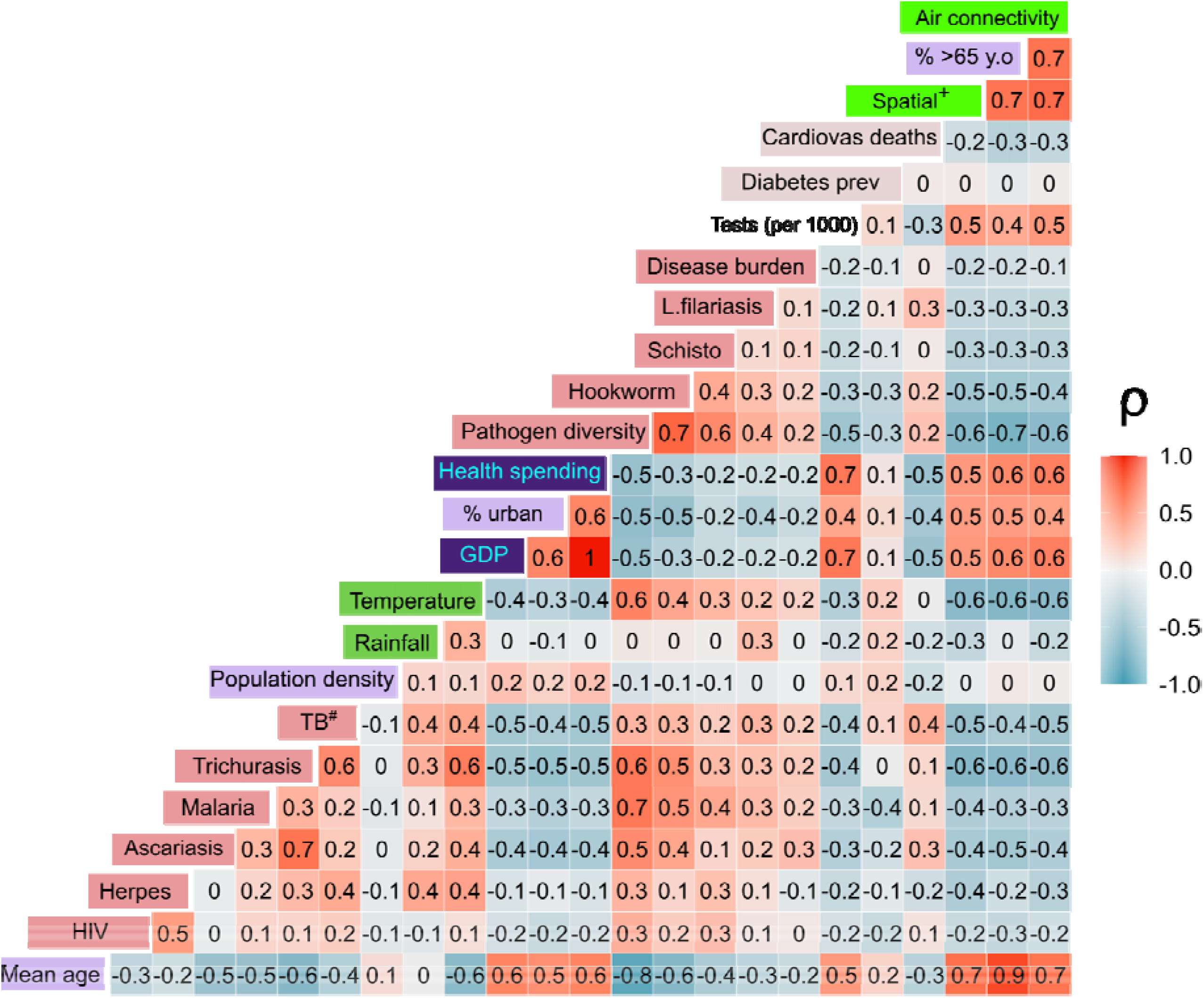
Pearson correlation heat map of the correlations between predictors. Rho values (ρ) are rounded up to 1 decimal place. Variables where ρ > 0.7 and had higher overall mean ρ values across all other variables were removed from the analysis.

**Table S2:** Leave-one-out (LOO) model comparison results for both models. Model performance is quantified using expected log pointwise predictive density (ELPD), expressed here as the relative estimates (ELPD) with respect to the best performing model. The performance of an alternative model is deemed comparable to the best model if the mean estimate lies within approximately one standard error of IZELPD = 0.

| Model name | mean ($\hat{\rho}$ELPD) | se of $\hat{\rho}$ELPD |
| --- | --- | --- |
| ***Cases Model*** |  |  |
| Reduced Set (all predictors) with region included in error term | 0.0 | 0.0 |
| Full case model (all predictors) region included in error term. | -1.9 | 6.5 |
| Full case model (all predictors) region not included in error term | -14.0 | 8.3 |
| ***Deaths Model*** |  |  |
| Reduced Set (all predictors) with region included in error term | 0.0 | 0.0 |
| Full case model (all predictors) region included in error term. | -10.9 | 4.3 |
| Full case model (all predictors) region not included in error term | -21.2 | 6.6 |

**Fig. S3:**
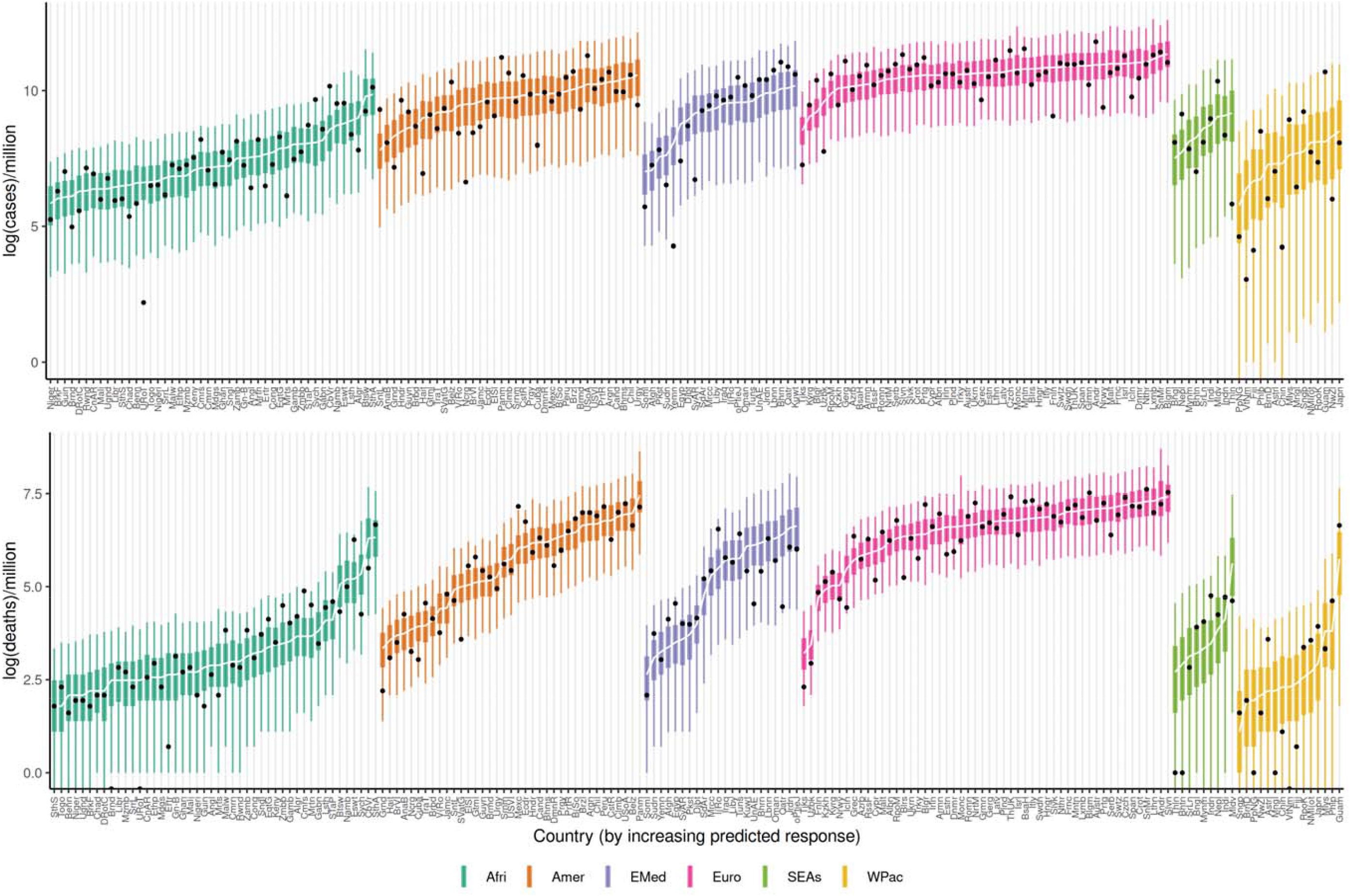
Predictive performance of our cases model (top panel) and deaths model (bottom panel) for each country. Countries are ordered by predicted values (lowest to highest). CIs are coloured by region. Afri: Africa, Amer = Americas, EMed = Eastern Mediterranean, SEAs = South East-Asia, WPac = Western Pacific. See Appendix S1 for country acronyms.

**Fig S4.**
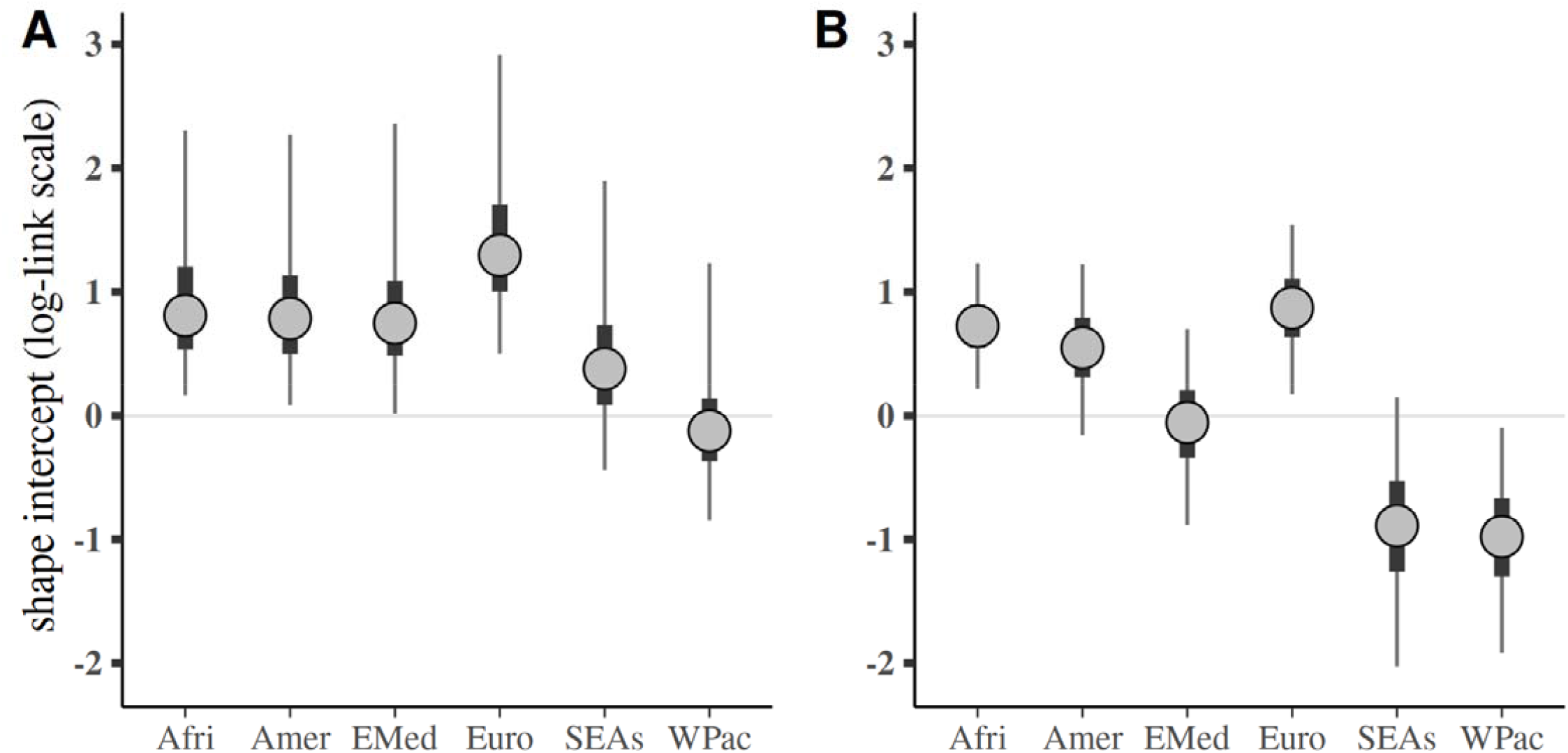
Region-level intercepts for the modelled overdispersion (shape) parameter: A) cases model; B) deaths models. For the negative binomial distribution, the variance is given by 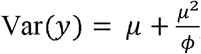, where *µ* is the mean and *log*(*ϕ*) is the shape parameter. The circles denote the posterior median and the thick and thin lines denote the 50% and 95% credible intervals, respectively. Afri = Africa, Amer = Americas, EMed = Eastern Medditeranean, SEAs = South-East Asia, WPac = Western Pacific.

